# Goal-Oriented Attentional Self-Regulation Training in Chronic Mild Traumatic Brain Injury Leads to Microstructural Plasticity in Prefrontal White Matter

**DOI:** 10.1101/2023.09.29.23296363

**Authors:** Haleh Karbasforoushan, Jamie Wren-Jarvis, Anna Hwang, Rachel Santiago, Sky Raptentsetsang, Lanya T. Cai, Jaclyn Xiao, Brian A. Maruyama, Gary M. Abrams, Tatjana Novakovic-Agopian, Pratik Mukherjee

## Abstract

Impaired attention is one of the most common, debilitating, and persistent consequence of traumatic brain injury (TBI), which impacts overall cognitive and executive functions in these patients. Previous neuroimaging studies, trying to understand the neural mechanism underlying attention impairment post TBI, have highlighted the role of prefrontal white matter tracts in attentional functioning in mild TBI. Goal-Oriented Attentional Self-Regulation (GOALS) is a cognitive rehabilitation training program that targets executive control functions in participants by applying mindfulness-based attention regulation and goal management strategies. GOALS training has been demonstrated to improve attention and executive functioning in patients with chronic TBI. However, its impact on microstructural integrity of attention-associated prefrontal white matter tracts is still unclear. Here, using diffusion MRI in a pilot randomized controlled trial, we investigated the effect of GOALS training on prefrontal white matter microstructure in US military veterans with chronic mild TBI (mTBI), compared to a matched control group of veterans with chronic mTBI who received standard of care brain health education. We also tested for an association between microstructural white matter changes and sustained attention ability in these patients pre- and post-GOALS training. Our results show significantly better white matter microstructural integrity in left and right anterior corona radiata in the GOALS group compared to the control group post-training. Moreover, we found a significant correlation between sustained attention ability of GOALS training participants and white matter integrity of their right anterior corona radiata pre- and post-training. Finally, our findings indicated that the improved white matter integrity of the anterior corona radiata in GOALS training participants was the result of increased neurite density and decreased fiber orientation dispersion within this tract.

## Introduction

Mild Traumatic Brain Injury (mTBI) is the most common type of TBI, counting for about 80% of all head trauma.^1,2^ Impaired attention is one of the most frequent, debilitating and persistent consequence of traumatic brain injury, which impacts overall cognitive and executive functions in these patients.^3–6^ Inability to sustain attention is the main contributor to poor memory, comprehending language, and executive control, so that it can impede rehabilitation of dysfunction in other cognitive domains as well.^3^ Cognitive rehabilitation in TBI has, therefore, been suggested to be largely focused on attention and executive functions training. ^7–9^

Brain structural changes in prefrontal cortex have been most frequently reported as the neural mechanism underlying the cognitive impairments post TBI.^10–12^ Moreover, using diffusion tensor imaging (DTI), we previously found the white matter fractional anisotropy (FA) of anterior corona radiata to be positively correlated with attentional functioning in chronic symptomatic mTBI and in controls.^13^ Anterior corona radiata (ACR) has also been shown to be the tract with most frequently low FA values in patients with chronic symptomatic mTBI, indicating impaired white matter microstructural integrity.^14^

Goal-Oriented Attentional Self-Regulation (GOALS) is a cognitive rehabilitation training program that targets executive control functions in participants by applying mindfulness-based attention regulation and goal management strategies.^15^ In contrast to training via practice on isolated tasks, this training protocol involves application of attention regulation skills and strategies to participant-defined goals in their own lives and community, in ecologically valid settings. One of the main training aims is to improve self-regulatory control mechanisms as they contribute to goal attainment. An overarching hypothesis is that improving attention regulation while engaging in goal-directed behavior will help establish more efficient and better integrated functional networks for the performance of goal-relevant tasks, and, ultimately, goal attainment in real-life contexts. GOALS training has been demonstrated to improve attention and executive functioning in patients with chronic TBI.^15–18^ However, it is still unclear if GOALS training could indeed lead to microstructural changes in attention-associated prefrontal tracts (e.g. anterior corona radiata), and whether these brain changes are correlated with attention improvements in the participants.

In this context, diffusion tensor imaging (DTI), a quantitative MRI technique, has provided important insights into white matter microstructure by providing information on the direction and degree of water diffusivity in white matter tracts.^19^ The degree of anisotropy (fractional anisotropy: FA) of water in the white matter tracts reflects their level of integrity.^20,21^ Evidence from human brain imaging studies suggests that white matter integrity could change with experience. For example, damage or degeneration to a tract is shown to be associated with a decrease in its white matter integrity (fractional anisotropy).^22–27^ Likewise, training-induced or experience-dependent increases in white matter integrity have also been reported in humans.^28-30^ Moreover, recently developed neurite orientation dispersion and density imaging (NODDI) methodology, based on biophysically inspired modeling of diffusion MRI data, provides further information about tissue microstructure, e.g., neurite density and fiber orientation dispersion.^31–34^ The neurite density index (NDI) is a measure of intracellular volume fraction and corresponds to axonal density in white matter; therefore higher NDI typically corresponds to better microstructural integrity. The fiber orientation dispersion index (ODI) is lowest in tracts with highly collimated white matter fibers; therefore, lower ODI typically corresponds to better microstructural integrity. NODDI has shown promise in the study of a range of disorders, such as neurodegenerative diseases,^35–38^ stroke,^39^ epilepsy,^40^ first-episode psychosis,^41^ amyotrophic lateral sclerosis (ALS),^42^ and mild TBI.^43,44^

To this end, we used diffusion MRI to examine the training-induced microstructural changes of prefrontal white matter tracts due to GOALS training in a group of US military veterans with chronic mTBI compared to a matched control group of veterans with chronic mTBI receiving standard of care brain health education (BHE). To our knowledge, this is the first randomized controlled trial to examine rehabilitation-related brain microstructural changes in a sample consisting exclusively of mTBI patients in the chronic phase. We tested our results via both region of interest (ROI) analysis, as well as voxel-wise tract-based spatial statistics (TBSS) analysis. Subsequently, the link between the microstructural integrity of prefrontal white matter tracts in individuals with GOALS training and their sustained attention scores pre- and post-training was tested using DTI. Finally, we used NODDI analysis to investigate whether better white matter integrity in the GOALS group compared to BHE group is the results of changes in tracts’ neurite density and orientation dispersion post-training.

## Materials and Methods

### Participants

This study was approved by Institutional Review Boards at the University of California San Francisco and San Francisco VA Medical Center. Participants were recruited from the San Francisco VA TBI Clinic, local VA community clinics, and Veteran groups using IRB approved information sheets and flyers. All participants provided informed consent before any study procedures. Inclusion criteria included: age of 18 or older; history of chronic mTBI (>6 months post-injury, sustained either in combat or as a civilian); stable psychoactive medication regimen; report of moderate to severe residual cognitive difficulties (via Neurobehavioral Symptom Inventory) that interfere with daily function; interest and availability to participate in cognitive training. History of mild TBI was confirmed through DOD/VA medical records and/or in-person Ohio State University TBI Instrument. Exclusion criteria included: history of moderate or severe TBI; unstable medical, neurological, or psychiatric conditions including psychosis, severe PTSD, severe anxiety, or depression precluding participation in research activities such as assessment and/or training; contraindications to MRI; illicit drug or alcohol use problems; or poor English comprehension. Following baseline evaluation, participants were placed in small groups, consisting of 2-3 participants of similar age, for training and the entire group was then randomized to receive either Goal-Oriented Attentional Self-Regulation (GOALS) or Brain Health Education (BHE) training.

### Neurocognitive Assessment of Sustained Attention

Participants were evaluated with a multi-level battery that included neuropsychological measures administered before and after GOALS or BHE trainings. Like our previous studies^45,46^, the current study used a neuropsychological battery specifically designed to assess Sustained Attention that is commonly affected by TBI and targeted by GOALS training. Assessments were administered by the same evaluator at both time points, and every attempt was made to administer them at the same time of the day. Evaluators were blinded to participants’ treatment conditions, and evaluators and therapists were separate individuals.

Sustained Attention was assessed using the Digit Vigilance Test (DVT) time and error scores^47^. The DVT requires sustained visual attention and accurate identification of target stimuli. Briefly, the participant was given a pencil and two pieces of paper, each containing rows of 35 single-digit numbers. The individual was instructed to cross out every “6” (or “9” post training, using alternate forms) as rapidly as possible. The examiner recorded the time in seconds that the individual took to complete both pages, as well as the number of errors of omission and commission. Both the time and the total error scores are included in the present normative system.

All neuropsychological raw test data were converted into standardized scores obtained from the corresponding normative manual which corrected for demographic factors^48^. These standardized scores were then transformed into z-scores for consistency. To assess the impact of training on targeted cognitive domains and reduce the number of multiple comparisons, z-scores for individual neuropsychological tests were averaged into the Sustained Attention scores.

### Interventions

GOALS and BHE were matched closely for time with therapists and training intensity. Both were administered across ten 2-hour group sessions, three 1-hour individual sessions, and 20 hours of home practice across 5 weeks. The interventions were conducted in a small group format with two to three participants and two therapists per group. Intervention manuals were written for instructors and participants.^49^

### GOALS training

GOALS training consisted of two key components: First, regulation of distractibility which is addressed via applied mindfulness-based attention regulation to redirect cognitive processes toward task-relevant activities. Participants learned to use a metacognitive strategy (‘‘Stop-Relax-Refocus’’) to stop activity when distracted, anxious, and/or overwhelmed; relax; and then refocus attention on the current primary goal; Second, active application of these goal-oriented attentional self-regulation skills to a range of situations and complex goals, from simple information processing tasks to challenging low-structure situations occurring in their own lives. Homework includes practice in maintaining goal-direction during challenging real-life situations identified by participants.

### BHE training

Brain-Health Education (BHE) training was an active comparison matched with GOALS for time with therapists, homework load, and group and individual session participation hours. It was also conducted in a small group format with two to five participants and two therapist per group. The BHE training was designed to be engaging and provide information about brain functioning and brain health. Although session materials included information about effects of stress, sleep, and diet, they were educational in nature, emphasizing knowledge and not skills. Group leaders did not assist participants with making connections between the material presented and possible positive effects on their own daily functioning, or how to integrate into their daily lives. Further, the presumed active ingredients of GOALS training, which include applied problem solving and attention regulation, were not part of the BHE intervention.

### Scanning Parameters

Participants underwent MRI acquisition at baseline and again post-intervention (approximately 6 weeks later) on a 3T Siemens Skyra scanner at the SF VA Medical Center with a 32-channel head coil. Anatomical MPRAGE scans were acquired using T1-weighting with the following parameters: repetition time (TR) = 2400ms; echo time (TE) = 2.24ms; flip angle = 8 degrees; in-plane resolution = 0.8mm; slice thickness = 0.8mm; number of slices = 208. Multi-shell multiband diffusion MRI was collected using the Human Connectome Project protocol consisting of spin-echo echo-planar imaging with the following parameters: multiband factor = 3, in-plane resolution = 1.8 mm, TR = 4550 ms, TE = 110 ms, flip angle = 90 degrees; matrix size = 118×118, FOV = 212 x 212 mm, slice thickness = 1.8mm, and number of slices = 78. Three diffusion MRI scans were collected with 71, 72 and 73 diffusion volumes in sequential order, each with intermixed b values of 0 s/mm^2^, 1500 s/mm^2^ and 3000 s/mm^2^.

### DTI Data Preprocessing

Each participant’s diffusion MRI data underwent quality control inspections and the same preprocessing pipeline to compute DTI and NODDI metrics. The FMRIB Software Library version 6.0.2 (FSL: Oxford Centre for Functional MRI of the Brain, UK; http://www.fmrib.ox.ac.uk/fsl/)^50,51^ was used for image preprocessing and DTI parameter computation. The three sequences with interleaved b=0 s/mm^2^, b=1500 s/mm^2^ and b=3000 s/mm^2^ shells were concatenated. A brain mask was created from the first volume of the multi-shell data using Freesurfer’s SynthStrip^52^. FSL’s *eddy* was applied to the raw multi-shell diffusion data to correct for motion and eddy current distortions, outlier replacement, susceptibility-by-movement, and slice-to-volume correction.^53–56^ A second brain mask was created from the first volume of the eddy corrected data and applied for skull stripping. The b=1500 s/mm^2^ shell volumes were extracted from the processed multi-shell data and used to calculate DTI parameters. To increase SNR, the b=0 s/mm^2^ were averaged together and used as the first volume followed by the 64 b=1500 s/mm^2^ volumes; this input was used in FSL’s *dtifit* to calculate fractional anisotropy (FA), mean diffusivity (MD), axial diffusivity (AD), and radial diffusivity (RD) maps. The full processed, multi-shell data including the b=3000 s/mm^2^ volumes was analyzed in the Accelerated Microstructure Imaging via Convex Optimization (AMICO) Toolbox^57^ to calculate the NODDI metric maps including neurite density index (NDI), orientation dispersion index (ODI) and free water fraction (FISO).

### Tract-Based Spatial Statistics Analysis

Voxel-wise statistical analysis of the DTI FA images was carried out using Tract-Based Spatial Statistics (TBSS^58^) in FSL. First, all subjects’ FA images were aligned to the standard 1×1×1 mm MNI152 template in FSL, using the non-linear registration tool (FNIRT). Next, the mean FA image was created and thinned to create a mean FA skeleton representing the centers of all tracts, using a threshold of 0.25. Each subject’s aligned FA map was then projected onto this skeleton resulting in each subject’s skeletonised FA image, and then masked by the prefrontal region template to limit the analysis to the prefrontal white matter.

Group differences in voxel-wise FA were examined by entering each subject’s prefrontal white matter skeletonised FA into a general linear model (two-sample t-test) design matrix with non-parametric permutation testing, using the Randomize tool in FSL (5000 permutations). The results were thresholded at p = 0.05 (corrected), using the threshold-free cluster enhancement (TFCE) option to find clusters without setting an initial cluster level^59^.

To further test whether sustained attention improvement in GOALS group participants was associated with their white matter integrity, we ran a within-group voxel-wise correlation analysis between GOALS participants’ prefrontal white matter skeletonized FA and their sustained attention scores pre- and post-training. Results were multiple comparisons corrected at p = 0.05 using the threshold-free cluster enhancement option to find the clusters with significant positive or negative correlation with sustained attention in GOALS group participants.

### Region-of-Interest (ROI) Analysis

The Johns Hopkins University (JHU) white matter atlas was used for identifying the regions of interest masks. We used the left and right anterior corona radiata as regions of interest for this study, as prior work demonstrated that this was the most frequently injured tract with abnormally low FA in chronic symptomatic mTBI^14^ and the one for which FA is correlated with attentional function^13^. The FA value within each ROI for each participant was calculated by taking the average voxel intensity of the skeletonized FA map within the binary mask of the ROI. Between-group comparisons for each ROI were then carried out by entering the individual subject FA value within the ROI into two-sample t-test analyses.

### Neurite Orientation Dispersion and Density Imaging (NODDI) Analysis

For NODDI ROI analysis, the NDI and ODI values within left and right anterior corona radiata for each participant was calculated by taking the average voxel intensity of the skeletonized NDI/ ODI map within the binary mask of the left and right anterior corona radiata. Also, for investigating the neurite density and orientation dispersion differences between groups within the cluster in which GOALS group had a higher white matter than BHE post-training, the cluster resulted from TBSS analysis was used as the region of analysis. Between-group comparisons for these ROIs were then carried out by entering the individual subject NDI and ODI values within the ROI into two-sample t-test analyses.

## Results

### Demographics

A total of 426 individuals were assessed for eligibility and 57 consented to this study. Of those who consented, 24 participants were later excluded from the study due to withdrawal or not being able to complete the training or post-training evaluations because of the COVID-19 shelter-in-place. Therefore, a total of 33 patients with mild TBI (19 in GOALS group and 14 in BHE group) completed the study procedures and were included in the present analyses (24% female; mean age=44.6 years [SD=14.2]; mean years education=15.2 [SD=2.35]). Head injuries were sustained from mixed causes including blunt force injuries, motor vehicle accidents, as well as blast waves. History of more than one mTBI (e.g., military combat training, martial arts training, high school football) was reported by 36% of participants. All participants were independent in basic activities of daily living but reported mild to moderate difficulties on tasks involving organization, problem solving, multitasking, and distractibility. Most participants were not working or going to school; 5 participants were gainfully employed and 10 were students. Demographics of two groups are presented in **Table 1**. There were no significant differences between the groups in demographic or clinical characteristics at baseline.

**Table 1.** Demographic Characteristics.

| Variable | GOALS |  | BHE |  | p |
| --- | --- | --- | --- | --- | --- |
| Number | 19 |  | 14 |  |  |
| Gender (% Female) | 21% |  | 29% |  | 0.42 |
| Race (% non-White) | 16% |  | 7% |  | 0.42 |
| Hispanic Ethnicity (%) | 15% |  | 14% |  | 0.79 |

|  | Mean | SD | Mean | SD | p |
| --- | --- | --- | --- | --- | --- |
| Age | 41.1 | 12.92 | 49.5 | 15.40 | 0.12 |
| Years of Education | 15.1 | 1.9 | 15.5 | 2.90 | 0.67 |
| Months since last injury | 118 | 98 | 176.3 | 191.00 | 0.31 |
| Mayo-Portland Total | 46.90 | 14.4 | 50.50 | 6.70 | 0.46 |
| BDI-II Total | 18.80 | 8.6 | 18.80 | 11.80 | 1.00 |
| PCL-M Total | 51.40 | 11.7 | 46.30 | 17.50 | 0.40 |
| Sustained Attention Z-score | -0.38 | 0.55 | -0.1 | 0.37 | 0.26 |

PCL-M = Post-Traumatic Stress Disorder Checklist-Military Version.
BDI-II = Beck Depression Inventory-II.
Sustained Attention represents the average of the group scores at baseline.
p-values provide the statistical GOALS vs. BHE comparison (Chi-square test for gender, race, and ethnicity, and two sample t-tests for all other variables.
Missing data: years of education, n=1 (GOALS); BDI-II and PCL-M, n=7 (GOALS), n=2 (BHE);
Sustained Attention score, n=1 (GOALS).

### Neurocognitive (Sustained Attention) Test Scores

There was no significant difference between the groups in sustained attention z-scores at the baseline (p = 0.26). However, a significant Group (GOALS vs. BHE) by Time (pre- vs. post-training) effect was identified for the Sustained Attention score (p = 0.03, F = 5.08), such that individuals who received GOALS demonstrated more improvement after treatment compared to those individuals who participated in the BHE intervention.

### Group Differences in White Matter Integrity based on ROI Analysis

Our ROI analysis of white matter tracts in prefrontal lobe (left and right anterior corona radiata, left and right superior corona radiata, and genu of corpus callosum) demonstrated no significant difference between GOALS and BHE groups before training. However, our ROI results revealed a significantly higher white matter FA in the GOALS group compared to BHE group post-training in left anterior corona radiata (t = 1.69, p = 0.034, GOALS: 0.52 +/- 0.02; BHE: 0.50 +/- 0.03) and right anterior corona radiata (t = 1.69, p = 0.041, GOALS: 0.51 +/- 0.02; BHE: 0.49 +/- 0.03).

We then tested for any association between these white matter differences in anterior corona radiata and the sustained attention measurement. Our ROI results indicated a moderate correlation between the sustained attention scores of the GOALS group participants pre- and post-training with their white matter integrity of right anterior corona radiata (p = 0.04, R = 0.32) (**Supplementary Figure 1A**). Moreover, when we tested for any association between the improvements in sustained attention scores and the changes in white matter integrity, we found that changes in white matter integrity of left anterior corona radiata in both groups pre- to post-training had a moderate correlation (R = 0.33) with the changes in their sustained attention scores at a strong trend level (p = 0.057) (**Supplementary Figure 1B**).

### Group Differences in White Matter Integrity Based on Voxel-wise Analysis

Similar to our results from ROI analysis, voxel-wise (TBSS) analysis indicated no significant difference between GOALS and BHE groups in white matter integrity of prefrontal lobe before training. The results of voxel-wise analysis indicated a significantly higher white matter FA in the GOALS group compared to BHE group post-training in left anterior corona radiata and left anterior thalamic radiation (**Figure 1A**). We also further explored the between groups voxel-wise differences post-training with a less conservative corrected p value < 0.1. As shown in **Figure 1B**, GOALS group had a higher white matter FA post-training in a number of left prefrontal white matter tracts, including left anterior corona radiata, left anterior thalamic radiation and left side of the genu of corpus callosum. There were no white matter tracts with lower FA in GOALS group than BHE group post-training even with a less conservative p value thereshold.

**Figure 1.**
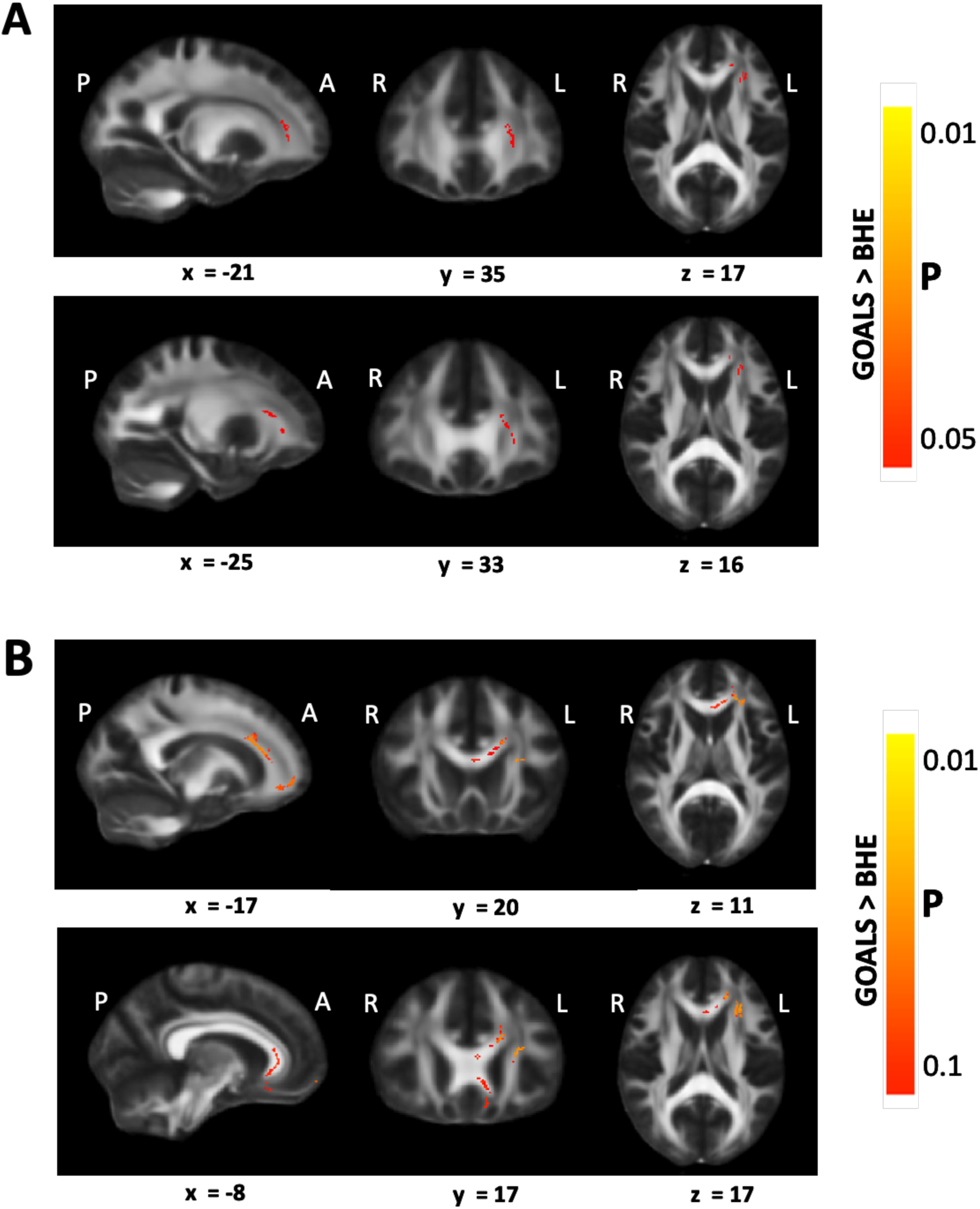
Tracts with higher white matter fractional anisotropy (FA) in GOALS group compared to BHE group post-training. (A) corrected p value < 0.05. Left anterior corona radiata showed significantly higher white matter FA in GOALS training participants compared to BHE training participants. (B) corrected p value <0.1. With a less conservative p value, left anterior corona radiata, left anterior thalamic radiation, and left side of the genu of the corpus callosum showed higher white matter FA in the GOALS group compared to BHE group. There was no tract with lower white matter integrity in the GOALS group compared to BHE group post-training even with a less conservative p value threshold.

We then tested for any correlation between white matter integrity of prefrontal tracts in the GOALS group with their sustained attention scores pre- and post-training using the same voxel-wise TBSS analysis. As shown in **Figure 2**, there was a significant positive correlation between white matter integrity of right anterior corona radiata and sustained attention scores in GOALS group pre- and post-training. There was no negative correlation between sustained attention scores and any prefrontal white matter tracts in the GOALS group.

**Figure 2.**
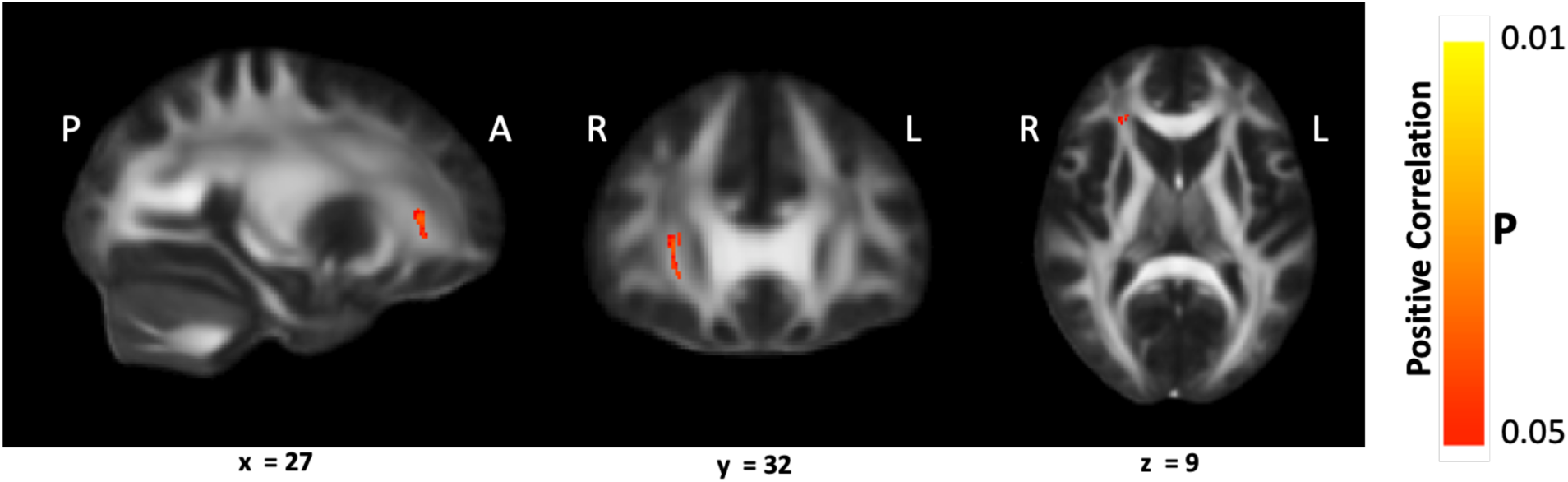
Tracts with significant correlation between white matter fractional anisotropy (FA) and sustained attention score in GOALS group. Sustained attention score in the GOALS group was positively correlated with the white matter FA of right anterior corona radiata pre- and post-training, after correction for multiple voxelwise comparisons (p < 0.05). There was no significant negative correlation between sustained attention scores and any white matter tracts in the GOALS group.

### Group Differences in White Matter Integrity Based on NODDI Analysis

We further investigated microstructural changes of the tracts in which the GOALS group had a higher white matter FA than BHE group post-training. On JHU ROI analysis, there was no significant between-group difference in neurite density index (NDI) or fiber orientation dispersion index (ODI) of left or right anterior corona radiata before training. However, there was a higher NDI in the GOALS group’s right anterior corona radiata than BHE group after training (p = 0.048, GOALS: 0.64 +/- 0.02; BHE: 0.61 +/- 0.06) with a strong trend towards higher NDI in the left anterior corona radiata as well (p = 0.057, GOALS: 0.66 +/- 0.03; BHE: 0.63 +/- 0.06). When testing the between-group differences in NDI and ODI post-training within the voxel cluster which had a significantly higher white matter FA in the GOALS group vs. BHE group, there was significantly higher NDI (p = 0.029, GOLAS: 0.63 +/- 0.03; BHE: 0.59 +/- 0.07) as well as significantly lower ODI (p = 0.005, GOALS: 0.16 +/- 0.01; BHE: 0.18 +/- 0.02) in the GOALS group than BHE group post-training.

## Discussion

Impaired attention is one of the most debilitating and persistent consequence of traumatic brain injury, and the main contributor to poor memory, comprehending language, and executive control, which can impede rehabilitation of dysfunction in other cognitive domains as well.^3–6^ Cognitive rehabilitation in TBI has, therefore, been suggested to be largely focused on attention and executive functioning training.^7–9^ Previous DTI studies, trying to understand the neural mechanism underlying attention impairment post TBI, have highlighted the role of prefrontal white matter tracts, especially decreased white matter integrity of anterior corona radiata in patients with mild TBI.^10–14^

Goal-Oriented Attentional Self-Regulation (GOALS) is a cognitive rehabilitation training program that targets executive control functions in participants by applying mindfulness-based attention regulation and goal management strategies.^15^ GOALS training improves attention and executive functioning in patients with chronic TBI.^15–18^ However, it has been still unclear if GOALS training could lead to microstructural changes in attention-associated prefrontal tracts (e.g. anterior corona radiata), and whether these brain changes are correlated with attention improvements in the participants.

In this study, we used DTI and NODDI in a proof-of-concept randomized controlled trial to investigate the effect of 5 weeks of GOALS training on the microstructural changes of prefrontal white matter in patients with chronic mild TBI, compared to a matched control mTBI group receiving standard of care. Moreover, we tested for any association between these microstructural changes and sustained attention ability in these patients pre- and post-GOALS training.

Our neurocognitive measurements indicated a significant group (GOALS vs. BHE) by Time (pre- vs. post-training) effect for the Sustained Attention score, such that individuals who received GOALS demonstrated more improvement after treatment compared to those individuals who participated in the BHE intervention. ROI analysis of white matter tracts from DTI in prefrontal lobe revealed a significantly higher white matter integrity in GOALS group compared to BHE group post-training in left and right anterior corona radiata tracts. Also, our voxel-wise TBSS analysis indicated a significant increased white matter integrity in GOALS group compared to BHE group post-training in left anterior corona radiata and left anterior thalamic radiation tracts. Moreover, from both ROI and voxel-wise correlation analyses, GOALS group participants showed a significant positive correlation between their sustained attention scores and the white matter integrity of right anterior corona radiata pre- and post-training.

Finally, NODDI analysis demonstrated a higher neurite density in the GOALS group’s left and right anterior corona radiata than the BHE group after training. When testing the between-group differences in NDI and ODI post-training within the voxel cluster which had a significantly higher white matter integrity in GOALS group vs. BHE group, we found that higher white matter FA in GOALS group, compared to BHE group, post-training was due to increased neurite density and decreased fiber orientation dispersion in this region of the anterior corona radiata. Therefore, rising intracellular volume fraction contributes to the observed neuroplasticity of anterior corona radiata on DTI, whereas improved fiber organization also plays a role in the core region of microstructural change within the anterior corona radiata.

These findings represent, to our knowledge, the first evidence of neuroplasticity associated with cognitive training in chronic mTBI. The attention training-related improvements of microstructural integrity in the anterior corona radiata are consistent with prior work implicating this prefrontal white matter tract in microstructural damage long-term post-mTBI^14^ and in attention deficits in chronic symptomatic mTBI patients.^13^ These proof-of-concept observations need to be validated in larger studies designed to assess the potential of diffusion MRI metrics as biomarkers for mTBI patient selection and for treatment response in trials of cognitive/behavioral interventions. Continuing progress in MR scanner hardware, pulse sequences and computational image processing, including deep learning and generative artificial intelligence, should yield further advances in the sensitivity and precision of diffusion MRI for brain microstructural plasticity as well as help translate this methodology into clinical practice.

### Transparency, Rigor and Reproducibility Statement

The study was pre-registered at clinicaltrials.gov (NCT02920788). The analysis plan was not formally preregistered. A sample size of 62 subjects was planned and the sample size for the present study is 33 subjects, 19 in the treatment group, and 14 in the active control group. Enrollment fell short of the target due to the COVID-19 pandemic. 426 potential participants were screened, imaging data were obtained from 43, and successfully analyzed in 33 who completed treatment, provided pre and post resting-state data, and met data quality assurance criteria. Imaging acquisition, imaging quality control decisions, and analyses were performed by team members blinded to treatment allocation of participants. All equipment and software used to perform imaging and preprocessing are widely available from commercial sources. The key inclusion criteria and outcome evaluations are established standards. Correction for multiple comparisons was performed using false discovery rate. At this time, no replication or external validation studies have been performed or are planned/ongoing to our knowledge, but our team is currently in the early stages of planning a replication and validation study. De-identified data from this study are not available in a public archive. De-identified data from this study will be made available (as allowable according to institutional IRB standards) by emailing the corresponding author. Analytic toolboxes used to conduct the analyses presented in this study are available as open-source software. The authors agree to provide the full content of the manuscript on request by contacting the corresponding author.

## Data Availability

All data produced in the present study are available upon reasonable request to the corresponding authors.

## Acknowledgments

We would like to acknowledge Jerry Chen, Marissa Cassar and Maria Kryza-Lacombe for valuable contributions to this project.

## References

1. Ruff R. Two decades of advances in understanding of mild traumatic brain injury. The Journal of head trauma rehabilitation 2005;20(1):5–18

2. Katz DI, Cohen SI, Alexander MP. Mild traumatic brain injury. Handbook of clinical neurology 2015;127(131-156

3. Stierwalt JA, Murray LL. Attention impairment following traumatic brain injury. Copyright© 2002 by Thieme Medical Publishers, Inc., 333 Seventh Avenue, New …: 2002.

4. Ashman TA, Gordon WA, Cantor JB, et al. Neurobehavioral consequences of traumatic brain injury. The Mount Sinai Journal of Medicine, New York 2006;73(7):999–1005

5. Shah SA, Goldin Y, Conte MM, et al. Executive attention deficits after traumatic brain injury reflect impaired recruitment of resources. Neuroimage: clinical 2017;14(233–241

6. Brenner LA. Neuropsychological and neuroimaging findings in traumatic brain injury and post-traumatic stress disorder. Dialogues in clinical neuroscience 2022;

7. Cicerone KD, Langenbahn DM, Braden C, et al. Evidence-based cognitive rehabilitation: updated review of the literature from 2003 through 2008. Archives of physical medicine and rehabilitation 2011;92(4):519–530

8. Cicerone KD, Goldin Y, Ganci K, et al. Evidence-based cognitive rehabilitation: systematic review of the literature from 2009 through 2014. Archives of physical medicine and rehabilitation 2019;100(8):1515–1533

9. Park NW, Ingles JL. Effectiveness of attention rehabilitation after an acquired brain injury: a meta-analysis. Neuropsychology 2001;15(2):199

10. Dall’Acqua P, Johannes S, Mica L, et al. Prefrontal cortical thickening after mild traumatic brain injury: a one-year magnetic resonance imaging study. Journal of Neurotrauma 2017;34(23):3270–3279

11. Levine B, Kovacevic N, Nica EI, et al. Quantified MRI and cognition in TBI with diffuse and focal damage. NeuroImage: Clinical 2013;2(534–541

12. Levine B, Kovacevic N, Nica E, et al. The Toronto traumatic brain injury study: injury severity and quantified MRI. Neurology 2008;70(10):771–778

13. Niogi SN, Mukherjee P, Ghajar J, et al. Structural dissociation of attentional control and memory in adults with and without mild traumatic brain injury. Brain 2008;131(12):3209–3221

14. Niogi S, Mukherjee P, Ghajar J, et al. Extent of microstructural white matter injury in postconcussive syndrome correlates with impaired cognitive reaction time: a 3T diffusion tensor imaging study of mild traumatic brain injury. American Journal of Neuroradiology 2008;29(5):967–973

15. Novakovic-Agopian T, Chen AJ-W, Rome S, et al. Rehabilitation of executive functioning with training in attention regulation applied to individually defined goals: a pilot study bridging theory, assessment, and treatment. The Journal of head trauma rehabilitation 2011;26(5):325–338

16. Novakovic-Agopian T, Kornblith E, Abrams G, et al. Training in goal-oriented attention self-regulation improves executive functioning in veterans with chronic traumatic brain injury. Journal of neurotrauma 2018;35(23):2784–2795

17. Novakovic-Agopian T, Posecion L, Kornblith E, et al. Goal-Oriented Attention Self-Regulation Training improves executive functioning in veterans with post-traumatic stress disorder and mild traumatic brain injury. Journal of Neurotrauma 2021;38(5):582–592

18. Novakovic-Agopian T, Kornblith E, Abrams G, et al. Long-term effects of executive function training among veterans with chronic TBI. Brain Injury 2019;33(12):1513–1521

19. Mukherjee P, Berman JI, Chung SW, et al. Diffusion tensor MR imaging and fiber tractography: theoretic underpinnings. AJNR Am J Neuroradiol 2008;29(4):632–41, doi:10.3174/ajnr.A1051

20. Pierpaoli C, Barnett A, Pajevic S, et al. Water diffusion changes in Wallerian degeneration and their dependence on white matter architecture. Neuroimage 2001;13(6):1174–1185, doi:10.1006/nimg.2001.0765

21. Virta A, Barnett AL, Pierpaoli C. Visualizing and characterizing white matter fiber structure and architecture in the human pyramidal tract using diffusion tensor MRI. Magnetic Resonance Imaging 1999;17(8):1121–1133, doi:Doi 10.1016/S0730-725x(99)00048-X

22. Werring DJ, Toosy AT, Clark CA, et al. Diffusion tensor imaging can detect and quantify corticospinal tract degeneration after stroke. Journal of Neurology, Neurosurgery & Psychiatry 2000;69(2):269–272

23. Thomalla G, Glauche V, Koch MA, et al. Diffusion tensor imaging detects early Wallerian degeneration of the pyramidal tract after ischemic stroke. Neuroimage 2004;22(4):1767–1774

24. Ward NS, Newton JM, Swayne OB, et al. Motor system activation after subcortical stroke depends on corticospinal system integrity. Brain 2006;129(3):809–819

25. Heise V, Filippini N, Ebmeier K, et al. The APOE ɛ4 allele modulates brain white matter integrity in healthy adults. Molecular psychiatry 2011;16(9):908

26. Kraus MF, Susmaras T, Caughlin BP, et al. White matter integrity and cognition in chronic traumatic brain injury: a diffusion tensor imaging study. Brain 2007;130(10):2508–2519

27. Vernooij MW, de Groot M, van der Lugt A, et al. White matter atrophy and lesion formation explain the loss of structural integrity of white matter in aging. Neuroimage 2008;43(3):470–477

28. Bengtsson SL, Nagy Z, Skare S, et al. Extensive piano practicing has regionally specific effects on white matter development. Nature neuroscience 2005;8(9):1148

29. Lövdén M, Bodammer NC, Kühn S, et al. Experience-dependent plasticity of white-matter microstructure extends into old age. Neuropsychologia 2010;48(13):3878–3883

30. Scholz J, Klein MC, Behrens TE, et al. Training induces changes in white-matter architecture. Nature neuroscience 2009;12(11):1370

31. Zhang H, Schneider T, Wheeler-Kingshott CA, et al. NODDI: practical in vivo neurite orientation dispersion and density imaging of the human brain. Neuroimage 2012;61(4):1000–1016

32. Jespersen SN, Kroenke CD, Østergaard L, et al. Modeling dendrite density from magnetic resonance diffusion measurements. Neuroimage 2007;34(4):1473–1486

33. Jespersen SN, Leigland LA, Cornea A, et al. Determination of axonal and dendritic orientation distributions within the developing cerebral cortex by diffusion tensor imaging. IEEE transactions on medical imaging 2011;31(1):16–32

34. Kamiya K, Hori M, Aoki S. NODDI in clinical research. Journal of Neuroscience Methods 2020;346(108908

35. Kamagata K, Hatano T, Okuzumi A, et al. Neurite orientation dispersion and density imaging in the substantia nigra in idiopathic Parkinson disease. European radiology 2016;26(2567–2577

36. Colgan N, Siow B, O’Callaghan JM, et al. Application of neurite orientation dispersion and density imaging (NODDI) to a tau pathology model of Alzheimer’s disease. Neuroimage 2016;125(739–744

37. Slattery CF, Zhang J, Paterson RW, et al. ApoE influences regional white-matter axonal density loss in Alzheimer’s disease. Neurobiology of aging 2017;57(8–17

38. Andica C, Kamagata K, Hatano T, et al. Neurite orientation dispersion and density imaging of the nigrostriatal pathway in Parkinson’s disease: Retrograde degeneration observed by tract-profile analysis. Parkinsonism & related disorders 2018;51(55–60

39. Adluru G, Gur Y, Anderson JS, et al. Assessment of white matter microstructure in stroke patients using NODDI. IEEE: 2014.

40. Sone D, Sato N, Ota M, et al. Abnormal neurite density and orientation dispersion in unilateral temporal lobe epilepsy detected by advanced diffusion imaging. NeuroImage: Clinical 2018;20(772–782

41. Rae CL, Davies G, Garfinkel SN, et al. Deficits in neurite density underlie white matter structure abnormalities in first-episode psychosis. Biological psychiatry 2017;82(10):716–725

42. Broad RJ, Gabel MC, Dowell NG, et al. Neurite orientation and dispersion density imaging (NODDI) detects cortical and corticospinal tract degeneration in ALS. Journal of Neurology, Neurosurgery & Psychiatry 2019;90(4):404–411

43. Palacios EM, Owen JP, Yuh EL, et al. The evolution of white matter microstructural changes after mild traumatic brain injury: a longitudinal DTI and NODDI study. Science advances 2020;6(32):eaaz6892

44. Yuh EL, Cooper SR, Mukherjee P, et al. Diffusion tensor imaging for outcome prediction in mild traumatic brain injury: a TRACK-TBI study. Journal of neurotrauma 2014;31(17):1457–1477

45. Novakovic-Agopian T, Chen AJ, Rome S, et al. Rehabilitation of executive functioning with training in attention regulation applied to individually defined goals: a pilot study bridging theory, assessment, and treatment. J Head Trauma Rehabil 2011;26(5):325–38, doi:10.1097/HTR.0b013e3181f1ead2

46. Novakovic-Agopian T, Kornblith E, Abrams G, et al. Training in Goal-Oriented Attention Self-Regulation Improves Executive Functioning in Veterans with Chronic Traumatic Brain Injury. J Neurotrauma 2018;35(23):2784–2795, doi:10.1089/neu.2017.5529

47. Lewis RF. Digit vigilance test. Psychological Assessment Resources: 1995.

48. Heaton RK. Revised comprehensive norms for an expanded Halstead-Reitan Battery: Demographically adjusted neuropsychological norms for African American and Caucasian adults, professional manual. Psychological Assessment Resources: 2004.

49. Binder D, Turner G, Chen A, et al. The brain health education workshop: Instructor and participant training manuals. San Francisco 2009;

50. Behrens TE, Woolrich MW, Jenkinson M, et al. Characterization and propagation of uncertainty in diffusion-weighted MR imaging. Magn Reson Med 2003;50(5):1077–88, doi:10.1002/mrm.10609

51. Smith SM, Jenkinson M, Woolrich MW, et al. Advances in functional and structural MR image analysis and implementation as FSL. Neuroimage 2004;23(S208–S219, doi:10.1016/j.neuroimage.2004.07.051

52. Hoopes A, Mora JS, Dalca AV, et al. SynthStrip: skull-stripping for any brain image. NeuroImage 2022;260(119474

53. Andersson JL, Graham MS, Zsoldos E, et al. Incorporating outlier detection and replacement into a non-parametric framework for movement and distortion correction of diffusion MR images. Neuroimage 2016;141(556–572

54. Andersson JL, Sotiropoulos SN. An integrated approach to correction for off-resonance effects and subject movement in diffusion MR imaging. Neuroimage 2016;125(1063–1078

55. Andersson JL, Graham MS, Drobnjak I, et al. Towards a comprehensive framework for movement and distortion correction of diffusion MR images: Within volume movement. Neuroimage 2017;152(450–466

56. Andersson JL, Skare S, Ashburner J. How to correct susceptibility distortions in spin-echo echo-planar images: application to diffusion tensor imaging. Neuroimage 2003;20(2):870–888

57. Daducci A, Canales-Rodríguez EJ, Zhang H, et al. Accelerated microstructure imaging via convex optimization (AMICO) from diffusion MRI data. Neuroimage 2015;105(32–44

58. Smith SM, Jenkinson M, Johansen-Berg H, et al. Tract-based spatial statistics: voxelwise analysis of multi-subject diffusion data. Neuroimage 2006;31(4):1487–505, doi:10.1016/j.neuroimage.2006.02.024

59. Smith SM, Nichols TE. Threshold-free cluster enhancement: addressing problems of smoothing, threshold dependence and localisation in cluster inference. Neuroimage 2009;44(1):83–98

