## Supplemental Figure 1 for "Goal-Oriented Attentional Self-Regulation Training in Chronic Mild Traumatic Brain Injury Leads to Microstructural Plasticity in Prefrontal White Matter"

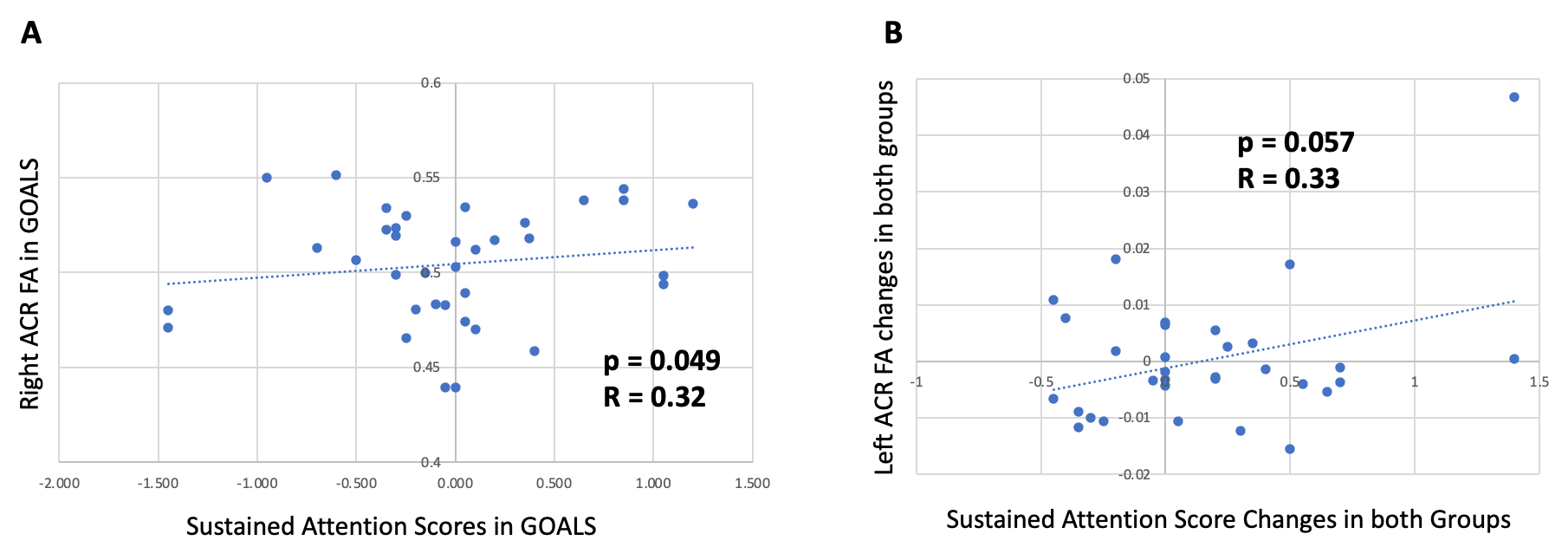

**Supplementary Figure 1. Correlation between white matter FA and Sustained Attention Scores. (A)** Correlation between the sustained attention scores and white matter FA of right anterior corona radiata in the GOALS group participants pre- and post-training. **(B)** Correlation between the improvements in sustained attention scores and the changes in white matter integrity of left anterior corona radiata in both groups pre- to post-training.
